# Safety and efficacy of the mRNA BNT162b2 vaccine against SARS-CoV-2 in five groups of immunocompromised patients and healthy controls in a prospective open-label clinical trial

**DOI:** 10.1101/2021.09.07.21263206

**Authors:** Peter Bergman, Ola Blennow, Lotta Hansson, Stephan Mielke, Piotr Nowak, Puran Chen, Gunnar Söderdahl, Anders Österborg, C. I. Edvard Smith, David Wullimann, Jan Vesterbacka, Gustaf Lindgren, Lisa Blixt, Gustav Friman, Emilie Wahren-Borgström, Anna Nordlander, Angelica Cuapio Gomez, Mira Akber, Davide Valentini, Anna-Carin Norlin, Anders Thalme, Gordana Bogdanovic, Sandra Muschiol, Peter Nilsson, Sophia Hober, Karin Loré, Margaret Sällberg Chen, Marcus Buggert, Hans-Gustaf Ljunggren, Per Ljungman, Soo Aleman, the COVAXID-collaborator group (shown separately)

## Abstract

**Background:** Patients with immunocompromised disorders have mainly been excluded from clinical trials of vaccination against COVID-19. Thus, the aim of this prospective clinical trial was to investigate the safety and efficacy after two doses of BNT162b2 mRNA vaccination in five selected groups of immunocompromised patients and healthy controls.

**Methods:** 539 study subjects (449 patients and 90 controls) were included in the clinical trial. The patients had either primary (n=90), or secondary immunodeficiency disorders due to human immunodeficiency virus infection (n=90), allogeneic hematopoietic stem cell transplantation/chimeric antigen receptor T cell therapy (n=90), solid organ transplantation (SOT) (n=89), or chronic lymphocytic leukemia (CLL) (n=90). The primary endpoint was seroconversion rate two weeks after the second dose. The secondary endpoints were safety and documented SARS-CoV-2 infection.

**Findings:** Adverse events were generally mild, but one case of fatal suspected unexpected serious adverse reaction occurred. 72·2% of the immunocompromised patients seroconverted compared to 100% of the controls (p=0.004). Lowest seroconversion rates were found in the SOT (43·4%) and CLL (63·3%) patient groups with observed negative impact of treatment with mycophenolate mofetil and ibrutinib, respectively.

**Interpretation:** The results showed that the mRNA BNT162b2 vaccine was safe in immunocompromised patients. The rate of seroconversion was substantially lower than in healthy controls, with a wide range of rates and antibody titres among predefined patient groups and subgroups. This clinical trial highlights the need for additional vaccine doses in certain immunocompromised patient groups and/or subgroups to improve immunity.

**Funding:** Knut and Alice Wallenberg Foundation, Nordstjernan AB, Region Stockholm, Swedish Research Council, Karolinska Institutet, and organizations for PID/CLL-patients in Sweden.

---

Coronavirus disease 2019 (COVID-19) was declared a pandemic by the World Health Organization (WHO) in March 2020. Immunocompromised patients were recognized early on in the pandemic as a high-risk group for severe disease with high rates of mortality ^1-3^. There are currently two approved mRNA vaccines, showing a good safety profile and high vaccine efficacy of 94-95% with regards to prevention of SARS-CoV-2 infection ^4,5^. Immunocompromised patients were not included in the pivotal trials. Thus, there is an unmet need for a clinical trial in which efficacy and safety data are prospectively evaluated in these vulnerable patient groups. The safety profile could be different due to elicitation of immune activation phenomena such as rejection of organ grafts or induction of graft-vs-host disease (GvHD) after allogeneic hematopoietic stem cell transplantation (HSCT). Emerging reports from cohort studies have also indicated poor antibody responses after COVID-19 vaccination in some immunocompromised patient groups ^6-10^. The aim of this clinical trial was to investigate safety and efficacy defined as the rate of seroconversion after two doses of BNT162b2 mRNA vaccine in five selected groups of immunocompromised patients compared to healthy controls.

## Methods

### Study design and participants

We conducted an open-label, non-randomized prospective clinical trial, in which the safety and efficacy of two doses of the mRNA BNT162b2 (Comirnaty^®^, Pfizer/BioNTech) vaccine were assessed in immunocompromised patients and healthy controls at the Karolinska University Hospital, Stockholm, Sweden. The sponsor of the study was Karolinska University Hospital. The study was approved by the Swedish Medical Product Agency (ID 5.1-2021-5881) and the Swedish Ethical Review Authority (ID 2021-00451). All participants provided written informed consent. This trial was registered at EudraCT (no. 2021-000175-37) and clinicaltrials.gov (no. 2021-000175-37). A description of the current trial with protocol is available via SciLifeLab Data Repository with the following doi: 10.17044/scilifelab.15059364 (English version) and 10.17044/scilifelab.15059355 (Swedish version).

Briefly, eligible for inclusion in the study were individuals ≥ 18 years of age, with no known history of SARS-CoV-2 infection who had either primary immunodeficiency disorders (PID) (n=90), or secondary immunodeficiency disorders due to infection with human immunodeficiency virus (HIV) (n=90), HSCT/chimeric antigen receptor T (CAR T) cell therapy (n=90), solid organ transplantation (SOT) (n=89), or chronic lymphocytic leukemia (CLL) (n=90). The control group (n=90) consisted of individuals without an immunocompromised disorder or treatment, and without significant co-morbidity. The controls were selected to represent three age groups each of which included 30 healthy individuals (18-39 years, 40-59 years, and >60 years, respectively). Exclusion criteria for the study were known diagnosis of previous or ongoing infection with SARS-CoV-2 assessed through patient interviews. Serology or PCR was not performed during screening (see further Procedures). Other exclusion criteria were coagulation disorder or treatment with anticoagulants which according to the investigator’s judgement contradicted an intramuscular injection; pregnancy or breastfeeding; history of an adverse reaction against the active substance or any of the components in the vaccine; incapability of giving informed consent or for another reason should be excluded according to the investigator’s judgement. The latter included clinical parameters such as the state of the underlying immunosuppressed disorder; e.g., ongoing rejection, infection, or severe GvHD. Furthermore, other vaccines planned to be given within 14 days before the first vaccine dose to 14 days after the second dose had to be postponed. The number of screened and included study subjects is shown in Figure 1. The main reasons for screening failure were previous COVID-19 infection, patient refusal, and that some study subjects already had been vaccinated outside the study. Detailed patient characteristics are outlined in Table 1.

**Figure 1.**
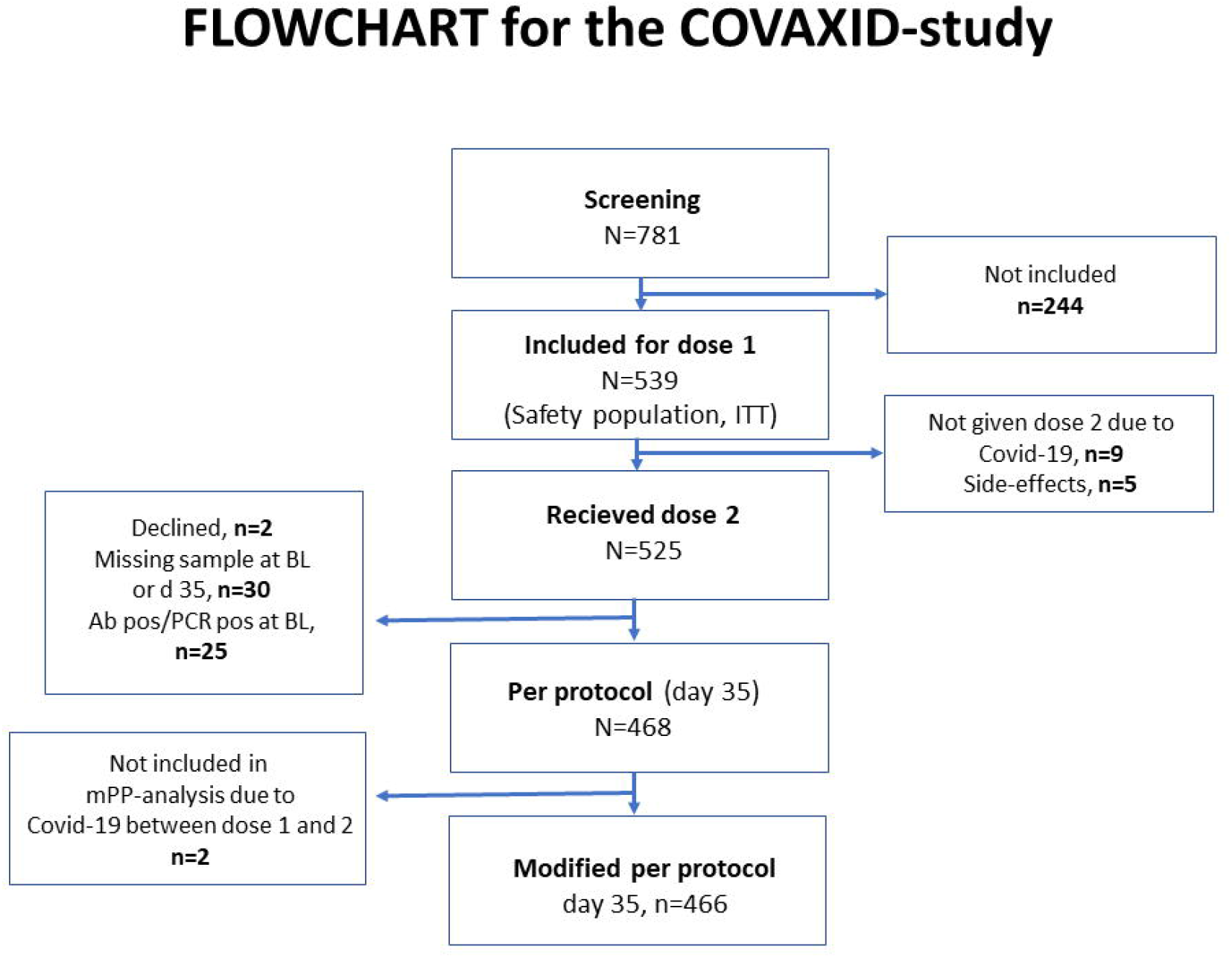
Flowchart of the study. The chart depicts the groups of study subjects screened prior to the study and the specific groups being enrolled and studied.

**Table 1:** Patient characteristics at baseline.

|  | **Controls**<br>**(n=90)** | **All**<br>**immunocompromised**<br>**(n=449)** | **PID**<br>**(n=90)** | **HIV**<br>**(n=90)** | **HSCT**<br>**(n=90)** | **SOT**<br>**(n=89<sup>1</sup>)** | **CLL**<br>**(n=90)** |
| --- | --- | --- | --- | --- | --- | --- | --- |
| **Sex, n (%)** |  |  |  |  |  |  |  |
| **Men** | 39 (43%) | 242 (54%) | 35 (39%) | 54 (60%) | 48 (53%) | 45 (51%) | 60 (67%) |
| **Women** | 51 (57%) | 207 (46%) | 55 (61%) | 36 (40%) | 42 (47%) | 44(49%) | 30 (33%) |
| **Age <65 years, n (%)** | 63 (70%) | 268 (60%) | 77 (86%) | 71 (79%) | 67 (74%) | 25 (28%) | 28 (31%) |
| **Laboratory parameters at baseline,**<br>**median (range)** |  |  |  |  |  |  |  |
| **IgG (g/L)** | 11·0 (7·2-21·2) (n=89) | 9·9 (1·0-34·4) (n=445) | 10·1 (3·5-26·1) | 12·8 (7·2-34·4) | 9·6 (1·6-17·9) (n=89) | 9·1 (3·4-25·0) (n=86) | 6·7 (1·0-20·8) |
| **Absolute lymphocyte count (x10<sup>9</sup>/L)** | 1·8 (1·0-4·1) (n=87) | 1·6 (0·2-112·6) (n=446) | 1·3 (0·4-9·6) | 1·8 (0·6-3·4) (n=89) | 1·6 (0·3-7·0) | 1·2 (0·2-2·8) (n=87) | 5·9 (0·4-112·6) |
| **Ongoing immunosuppression, n (%)** |  |  |  |  |  |  |  |
| **Corticosteroids** | 0 | 26 (6%) | 12 (13%) | 1 (1%) | 13 (14%) | 82 (92%) | 0 |
| **Other immunosuppressive agents** | 0 | 159 (35%) | 13 (14%) | 0 | 27 (30%) | 89 (100%) | 30 (33%)* |
| **Subgroups, (n)** | 1. 18-39 years (n=30)<br>2. 40-59 years (n=30)<br>3. >60 years (n=30) |  | 1. CVID (n= 50)<br>2. XLA (n=4)<br>3. Low number or defect<br>T-cell function (n=14)<br>4. Monogenic diseases<br>(n=10)<br>5. Other with expected<br>normal response (n=12) | 1. Latest CD4 T cell<br>count ≤ 300 cells/ul<br>(n= 30)<br>2. Latest CD4 T cell<br>count >300 cells/ul<br>(n= 60) | 1. CAR T (n=3), Allo<br>HSCT (n= 87)<br>Time after allo HSCT:<br>2. Early <6 mo (n=10)<br>3. Intermediate 6 - 12 mo<br>(n=12)<br>4. Late >12 mo (n=65) | Time after<br>transplantation:<br>1. ≤6 mo (n=33)<br>with/without MMF<br>2. >6 mo with MMF<br>(n=20)<br>3. >6 mo without<br>MMF (n=36) | 1. Indolent<br>untreated (n=30)<br>2. Ongoing<br>treatment with<br>ibrutinib (>6 mo)<br>(n=30)<br>3. Previous ibrutinib<br>treatment (≥2 mo<br>ago), now in off-<br>phase (n=10)<br>4. Previous treatment<br>with CD20 mAb<br>(>6 mo - <30 mo)<br>(n=20) |

**Abbreviations:** n: number, PID: primary immunodeficiency disorders, HIV: human immunodeficiency virus, HSCT: hematopoietic stem cell transplantation, SOT: solid organ transplantation, CLL: chronic lymphocytic leukemia, IgG: immunoglobulin G, CVID: common variable immunodeficiency, XLA: X-linked agammaglobulinemia, CD: cluster of differentiation, CAR T: chimeric antigen receptor T-cell therapy, mo: months, MMF: mycophenolate mofetil, mAb: monoclonal antibody, BL: baseline.
**Footnote:** \*all n=30 were on ibrutinib. <sup>1</sup>The different transplants in the SOT-group (n=89) were: 57 liver, 26 kidney, 6 kidney and pancreas.

### Procedures

The participants were given injections of BNT162b2 mRNA vaccine in standard dose (30 micrograms) into the deltoid muscle of the non-dominant arm on days 0 and 21 of the study; i.e., in a two-dose regimen according to the label. All vaccine doses were derived from the same batch (batch number EP2163). Blood samples were taken at day 0 (before the first vaccination), and then at days 10, 21 (before the second vaccination), and 35 (analysis of the primary endpoint). Serum samples were analyzed using quantitative test Elecsys^®^ Anti-SARS-CoV-2 S (Roche Diagnostics) on the Cobas 8000 e801pro for detection of antibodies to SARS-CoV-2 spike protein receptor binding domain (RBD). The measuring range is between 0·40 and 250 U/mL with cut-off for positive results at ≥ 0·80 U/mL. Positive samples with antibody titres of >250 U/mL were re-tested following a 1/10 dilution, and in applicable cases also a 1/100 dilution which increased the upper level of measuring range to 25,000 U/mL. Nasopharyngeal SARS-CoV-2 swab tests for real-time RT-PCR were taken before vaccination at day 0, and in case of symptoms of possible COVID-19 during follow-up. Hematological and biochemical assays were performed at days 0, 21, and 35. Study data including baseline characteristics, assay results, reactogenicity, adverse events, and concomitant medications were recorded in an electronic case report form (eCRF).

### Outcomes

The primary endpoint definition was seroconversion to the SARS-CoV-2 spike glycoprotein 14 days (day 35) after the second dose of vaccine in the per protocol (PP) population (n=468), being seronegative at study entry and who received two doses of vaccine (Figure 1). A PP (n=468) as well as a modified per protocol (mPP) population (n=466) were analyzed. The mPP excluded two patients who developed COVID-19 between study entry and day 35 (see Figure 1). The main secondary endpoint was safety and tolerability of the vaccine. This was analyzed on all patients receiving at least one dose of vaccine (safety population; intention to treat (ITT) population) (see Figure 1). An additional secondary endpoint was occurrence of SARS-CoV-2 infection with assessment of severity ^11^ (Supplementary Text).

### Safety and tolerability assessments

Reactogenicity was assessed by recording specific local (pain, erythema, or swelling at injection site) or systemic (fever, chill, headache, tiredness/fatigue, diarrhea, vomiting, new/worsened muscle- or joint pain) side effects as reported by patients in a paper diary for seven days following each vaccine dose. All reactogenicity events were graded as none/mild (grade 0-1), moderate (grade 2), severe (grade 3), life-threatening (grade 4), or death (grade 5) according to the Common Terminology Criteria for Adverse Events (CTCAE) (Supplementary Table 1). Other, non-reactogenicity associated adverse events (AE) were recorded until 14 days after administration of the second dose by patient interviews in conjunction with the second dose (day 21) and through a phone call two weeks following the 2^nd^ dose. Severe adverse events (SAE) and suspected, unexpected, serious adverse reactions (SUSAR) were assessed and recorded from the first vaccine dose to 6 weeks after the second dose, with exception of events related to the expected course of the main underlying disease.

### Statistical analysis

At the time of the study design, no information existed regarding the expected seroconversion rate of immunosuppressed individuals following vaccination with the mRNA BNT162b2 vaccine. Based on the initial BNT162b2 vaccine clinical trials results, we hypothesized that the proportion of seroconversion in healthy controls would be 99%. Choosing a sample size n=90 per group would give a power value of 81%, even with a conservatively low expected 10% difference in seroconversion in immunocompromised groups versus healthy controls. Analyses of the primary efficacy endpoint with seroconversion was performed on both the PP and the mPP populations, with estimation of the proportion of participants with seroconversion (95% confidence interval, CI). Comparisons of reactogenicity events between patient groups and controls were performed with Fisher’s exact test. Proportions of seroconversion were compared in patient groups, or prespecified subgroups *vs*. controls, with estimation of CIs and p-values (Fisher’s exact test). Logistic regression, univariable or multivariable, was used to analyze possible predictive factors for seroconversion failure. P values <0·05 were considered statistically significant. The statistical analyses were performed using R base (R Core Team, 2021). Additional details of statistical analyses are described in Supplementary Text.

## Results

### Participants

781 individuals were screened for eligibility for the study between February 12^th^ and February 22^nd^, 2021. Of these, 539 individuals were included in the trial (safety population; intention to treat (ITT)) (Figure 1). Each of the five patient groups and the control group consisted of 90 patients, with the exception of the SOT group (89 patients). All 539 included patients received the first dose of vaccine between February 23^rd^ and March 30^th^, 2021. Baseline characteristics of the ITT group is described in Table 1. All but fourteen (2·6%) study subjects went on to the second dose (Figure 1). Those that did not receive the second dose were study subjects diagnosed with COVID-19 (n=9) or that got side effects that prevented further vaccination (n=5) (Figure 1).

### Safety

#### Reactogenicity

Local and systemic reactogenicities, as reported by the study subjects in diaries, are presented in Supplementary Table 1. The proportions of patients and controls reporting reactogenicity events were not markedly different from each other in an overall comparison. However, a somewhat higher rate of systemic reactogenicity events was observed in the healthy control group than in the patient group (p<0.01) following the second dose, possibly due to some of the patient’s immunosuppressed status.

#### Adverse events

Other non-reactogenicity related AE, as reported by the study subjects by physical visits and telephone interviews are presented in Supplementary Table 2. A higher number of non-reactogenicity related AEs were registered in the patient groups compared to the controls regarding total numbers, grades 2-4 CTCAE, and these were possibly/probably related to the vaccine (Supplementary Table 2). Most AEs were from allogeneic HSCT/CAR T cell treated patients (n = 50), followed by patients with PID (n = 36), and SOT patients (n = 26). The most frequently reported AEs were infections; all assessed as unlikely to be related to the vaccine. Notably, two patients having undergone HSCT had activation of GvHD with altered liver function tests that required treatment with corticosteroids and consequently did not proceed to the second dose. Two additional patients, among those who received two doses, developed chronic GVHD of the skin and signs of obliterative bronchiolitis with worsened respiratory dysfunction after discontinuing immunosuppression before the first dose of vaccine, respectively. Finally, three patients developed CTCAE grade 2 cytopenias (thrombocytopenia n=1; neutropenia n=2), which were self-resolving without intervention (Supplementary Table 2).

#### Severe Adverse Events (SAE) and Severe Unexpected Serious Adverse Reaction (SUSAR)

Twenty-eight SAE were registered in a total of 24 patients during the study period (Table 2). Five SAE were assessed as possibly being linked to the vaccination, including (i) one vasovagal reaction in a HIV patient (moderate), (ii) febrile neutropenia in a HSCT patient (moderate), (iii) rejection in a liver transplanted patient (severe), and (iv) syncope in another liver transplanted patient (moderate). In addition, a SUSAR occurred in the HSCT-group. Five months after an allogeneic HSCT with prior CD19 CAR T treatment, the patient developed fever, vomiting, signs of disorientation, and respiratory distress four days after the first vaccination. This led to hospitalization and subsequent referral to the intensive care unit with suspicion of an immunologically driven pneumonia (bronchiolitis obliterans organizing pneumonia). No second vaccine dose was given. The patient responded well to corticosteroids and could be discharged after three weeks. Unfortunately, the patient later developed progressive diffuse pulmonary infiltrates resistant to broad anti-infectious and immunosuppressive treatment, and subsequently required ventilator therapy. The patient died two months after the first vaccination. An autopsy was performed revealing lung failure as the major cause of death. The case was assessed by the investigator and the sponsor to be likely related to the vaccination and has been reported as a SUSAR. Final results from both autopsy and additional immunological analyses are awaited and will be reported separately. Overall, the number of SAEs was highest in the SOT group and lowest in the people living with HIV (PLWH) group (below referred to as the HIV group). No SAE was observed in the healthy control group (Table 2).

**Table 2:** Severe adverse events (SAE) after two doses of BNT162b2 vaccine in healthy controls and five different groups of immunocompromised patients.

|  |  | **Controls**<br>**(n=90)** | **PID**<br>**(n=90)** | **HIV**<br>**(n=90)** | **HSCT**<br>**(n=90)** | **SOT**<br>**(n=89)** | **CLL**<br>**(n=90)** | **Total**<br>**(n=539)** |
| --- | --- | --- | --- | --- | --- | --- | --- | --- |
| **Events<sup>1</sup>** | SAE (events, n) | 0 | 3 | 2 | 5<sup>3</sup> | 12 | 6 | **28** |
|  | SAE (patients, n)<sup>1</sup> | 0 | 3 (3%) | 2 (2%) | 4 (4%)<sup>3</sup> | 12 (13%) | 3 (3%) | **24 (4%)** |
| **Related to vaccine<sup>2</sup>** | Possible (n, %) | 0 | 0% | 1 (50%) | 2 (40%)<sup>3</sup> | 2 (17%) | 0% | **5** |
|  | Unlikely (n, %) | 0 | 3 (100%) | 0% | 0% | 10 (83%) | 6 (100%) | **19** |
|  | Not related (n, %) | 0 | 0% | 1 (50%) | 3 (60%) | 0% | 0% | **4** |
| **Grading<sup>2</sup>** | Severe (n, %) | 0 | 1 (33%) | 1 (50%) | 3 (60%) | 4 (33%) | 0 | **9** |
|  | Moderate (n, %) | 0 | 1 (33%) | 1 (50%) | 2 (40%) | 8 (67%) | 6 (100%) | **18** |
|  | Mild (n, %) | 0 | 1 (33%) | 0% | 0% | 0% | 0% | **1** |
| **Resolved<sup>2</sup>** | Yes (n, %) | 0 | 3 (100%) | 1 (50%) | 5 (100%) | 6 (50%) | 5 (83%) | **20** |
|  | No (n, %) | 0 | 0% | 1 (50%) | 0% | 6 (50%) | 1(17%) | **8** |

**Abbreviations:** SAE: severe adverse reaction, PID: primary immunodeficiency, HIV: human immunodeficiency virus, HSCT: hematopoietic stem cell transplantation, SOT: solid organ transplantation, CLL: chronic lymphocytic leukemia.
**Footnotes:** <sup>1</sup>Percentage was calculated as the proportion of patients with at least one SAE in the patient-group.
<sup>2</sup>Percentage was calculated as the proportion of patients with at least one SAE divided by the total numbers of patients with at least one SAE. <sup>3</sup>One SUSAR occurred in this group.

### Primary endpoint: Seroconversion at day 35

The results of the PP analyses differed only marginally from the mPP analyses (Table 3 and Supplementary Table 3). Because of this, we chose to present the results from the mPP analyses. 466 study subjects (388 immunosuppressed patients in 5 groups and 78 healthy controls) were eligible for analyses (Figure 1). Results in terms of seroconversion and antibody titres from spike-specific IgG measurements are displayed in Figure 2 (patient group analyses) and in Figure 3 (patient subgroup analyses) as well as in Supplementary Figure 1 (patients group analyses including study subjects with SARS-CoV-2 antibody/PCR positivity at baseline). 72·2% of the patients in the mPP group seroconverted at day 35, compared to 100% of the controls (p=0.004) (Table 3). With exception of the HIV group, all patient groups showed a significantly higher likelihood for failure to seroconvert at day 35 compared to the control group. The highest seroconversion-failure rate was found in the SOT group, followed by the CLL group, PID group, HSCT group and the HIV group (Table 3 and Figure 2A).

**Table 3:** Numbers and proportions of seroconversion (modified per protocol; n=466) after two doses of BNT162b2 vaccine in healthy controls and five different groups of immunocompromised patients.^1^.

|  | Controls | All immunocompromised patients | PID | HIV | HSCT | SOT | CLL |
| --- | --- | --- | --- | --- | --- | --- | --- |
| **Seroconverted (n)** | 78 | 280 | 55 | 78 | 61 | 36 | 50 |
| **Seronegative (n)** | 0 | 108 | 20 | 1 | 11 | 47 | 29 |
| **Total (n)** | 78 | 388 | 75 | 79 | 72 | 83 | 79 |
| **Proportion of seroconverted (CI) (%), P-value** | 100<br>(95.4-100)<br>Ref. | 72.2<br>(67.4 – 76.6)<br>P<0.001 | 73.3<br>(61.9-82.9)<br>P<0.01 | 98.7<br>(93.1-100)<br>P=1 | 84.7<br>(74.3-92.1)<br>P<0.01 | 43.4<br>(32.5-54.7)<br>P<0.01 | 63.3<br>(51.7-73.9)<br>P<0.01 |

**Abbreviations:** PID: primary immunodeficiency, HIV: human immunodeficiency virus, HSCT, hematopoietic stem cell transplantation, SOT: solid organ transplantation, CLL: chronic lymphocytic leukemia, CTRL: healthy controls, CI: 95% confidence interval (estimated).
**Footnote:** <sup>1</sup>P-values of the differences vs. healthy controls were calculated.

**Figure 2.**
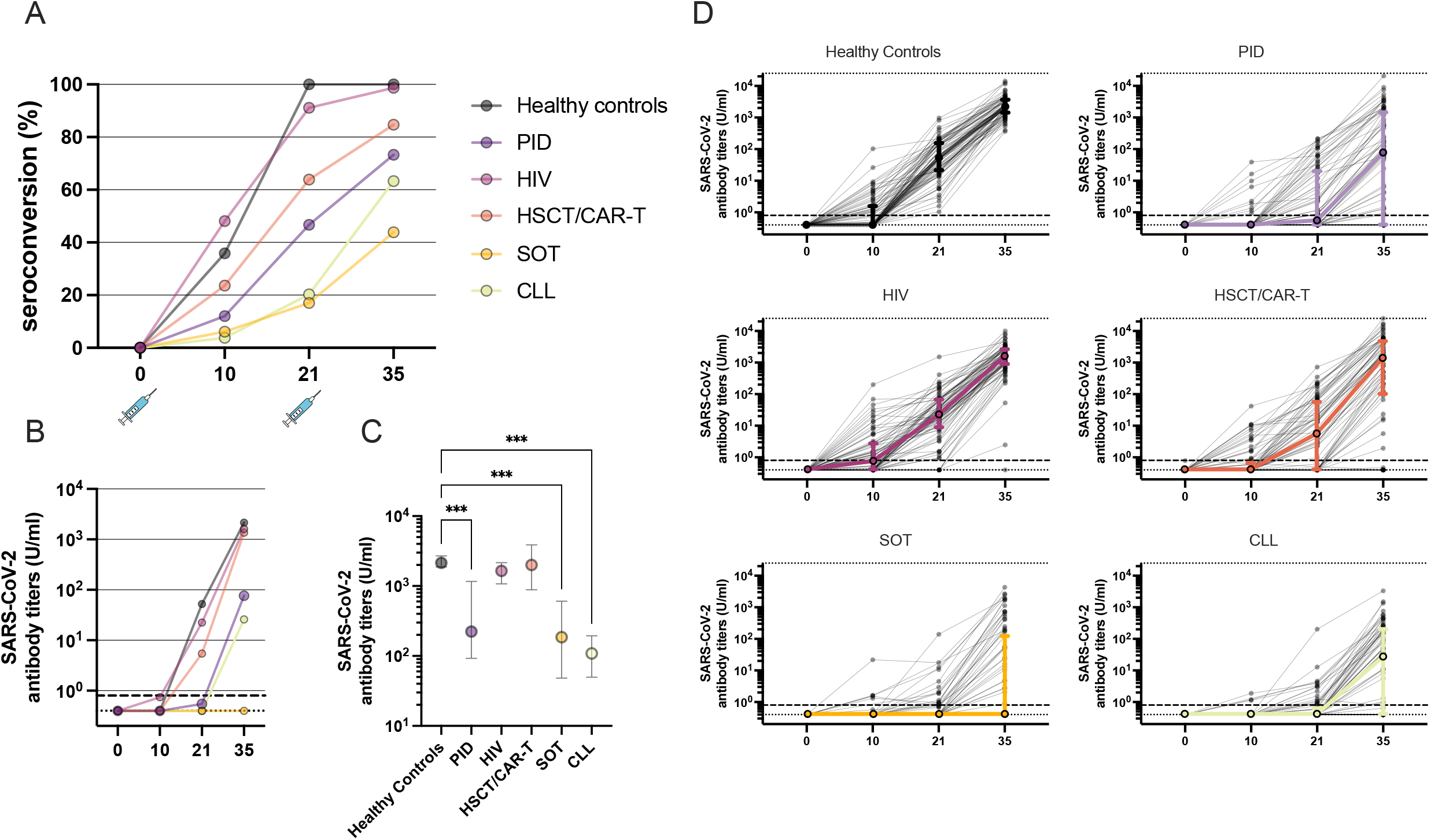
Subgroup analysis of seroconversion and antibody titers. A) Seroconversion analyzed according to modified per protocol (mPP) defined as ≥0.8 U/ml. B) Median SARS-CoV-2 specific antibody titers in the 6 cohorts. C) Median (CI 95%) SARS-CoV-2 specific antibody titers at day 35 in individuals who seroconverted in the 6 cohorts. For comparison, Kruskal-Wallis One-Way ANOVA and Dunn’s post hoc test was used. D) Individual antibody dynamics (black thin lines) with median (IQR) (thick lines) for each cohort.

Analyzing the different patient groups separately, the overall seroconversion rate in the SOT group was 43·4% (p<0.001 compared to controls). Analyzing the subgroups, patients receiving mycophenolate mofetil (MMF) had a significantly lower seroconversion rate than controls regardless of time after transplantation; 13·3% in patients <6 months after transplantation (p=0.01) and 10·0% in patients >6 months after transplantation (p<0.01). In contrast, the subgroup of patients not receiving MMF and vaccinated >6 months after transplantation had a seroconversion rate not differing significantly from controls (90·9% vs 100%, p=0.06) (Table 4 and Figure 3A). In multivariate analysis, MMF-treatment was an independent predictor for seroconversion failure (Table 5).

**Table 4:** Numbers and proportions of seroconversion for each patient group divided into subgroups.^1^.

|  | PID |  |  |  |  | HIV |  | HSCT |  |  |  | SOT |  |  | CLL |  |  |  |
| --- | --- | --- | --- | --- | --- | --- | --- | --- | --- | --- | --- | --- | --- | --- | --- | --- | --- | --- |
|  | CVID | XLA | CD4-cyt | Monog. Dis. | Other | >CD4 300 | <CD4 300 | CAR T | Early | Interm | Late | <6 mo | 6 mo MMF | 6 mo non-MMF | Indol | Previous CD20-mAb | Ibru | Off ibru |
| **Seropositive (n)** | 28 | 0 | 10 | 7 | 10 | 54 | 24 | 0 | 3 | 8 | 50 | 4 | 2 | 30 | 22 | 16 | 7 | 5 |
| **Seronegative (n)** | 13 | 4 | 1 | 2 | 0 | 0 | 1 | 2 | 1 | 4 | 4 | 26 | 18 | 3 | 4 | 2 | 19 | 4 |
| **Total (n)** | 41 | 4 | 11 | 9 | 10 | 54 | 25 | 2 | 4 | 12 | 54 | 30 | 20 | 33 | 26 | 18 | 26 | 9 |
| **Proportion of sero-converted (CI) (%)** | 68.3<br>(51.9-81.9)<br>P<0.01 | 0<br>(0-60.2)<br>P<0.01 | 90.9<br>(58.7-99.8)<br>P=0.12 | 77.8<br>(40-97.2)<br>P<0.01 | 100<br>(69.2-100)<br>P=1 | 100<br>(93.4-100)<br>P=1 | 96<br>(79.6-99.9)<br>P=0.24 | 0<br>(0-84.2)<br>P<0.01 | 75<br>(19.4-99.4)<br>P=0.05 | 66.7<br>(34.9-90.1)<br>P<0.01 | 92.6<br>(82.1-97.9)<br>P=0.03 | 13.3<br>(3.8-30.7)<br>P<0.01 | 10<br>(1.2-31.7)<br>P<0.01 | 90.9<br>(75.7-98.1)<br>P=0.02 | 84.6<br>(65.1-95.7)<br>P<0.01 | 88.9<br>(65.3-98.6)<br>P=0.03 | 26.9<br>(11.6-47.8)<br>P<0.01 | 55.6<br>(21.2-86.3)<br>P<0.01 |

**Abbreviations:** PID: primary immunodeficiency; CVID, common variable immunodeficiency; XLA, X-linked agammaglobulinemia, CD4-cyt: idiopathic CD4-cell lymphocytopenia, Monog. Dis: monogenic disorder, HIV: human immunodeficiency virus; CD4: CD4+ T-cells, HSCT, hematopoietic stem cell transplantation; CAR T, chimeric antigen receptor T-cells. Early, <6 months after transplantation; Interm, 6-12 months after transplantation; Late, >12 months after transplantation. SOT: solid organ transplantation; MMF, mycophenolate mofetil. CLL: chronic lymphocytic leukemia; Indol, indolent and not treated; Previous CD20-1b, previous treatment with BR/FCR bendamustine and rituximab / fludarabine, cyclophosphamide and rituximab; Ibru, ongoing ibrutinib treatment; Off ibru, off ibrutinib treatment for >2 months. CI: 95% confidence interval.
**Footnote:** <sup>1</sup>P-values of the differences vs. healthy controls were calculated.

**Figure 3.**
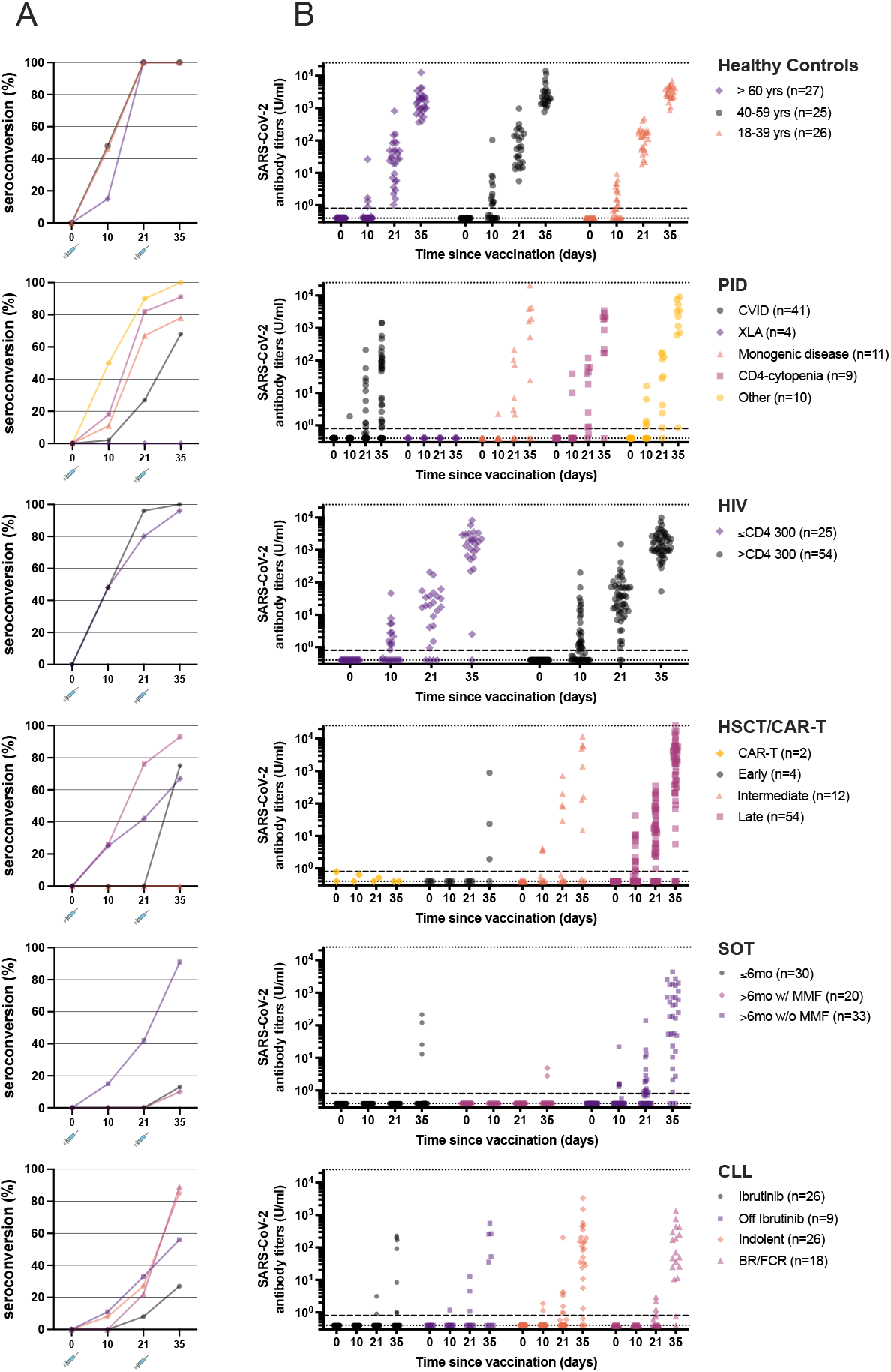
Subgroup analysis of seroconversion and antibody titers. A) Seroconversion analyzed according to modified per protocol (mPP) defined as ≥0.8 U/ml (see right legends for subgroup classification). B) Individual SARS-CoV-2 specific antibody titers for each timepoint in respective subgroup. Dotted lines represent upper (25 000 U/ml) and lower (0.4 U/ml) limits of detection. Dashed line represent seroconversion threashold 0.8 U/ ml.

**Table 5:**
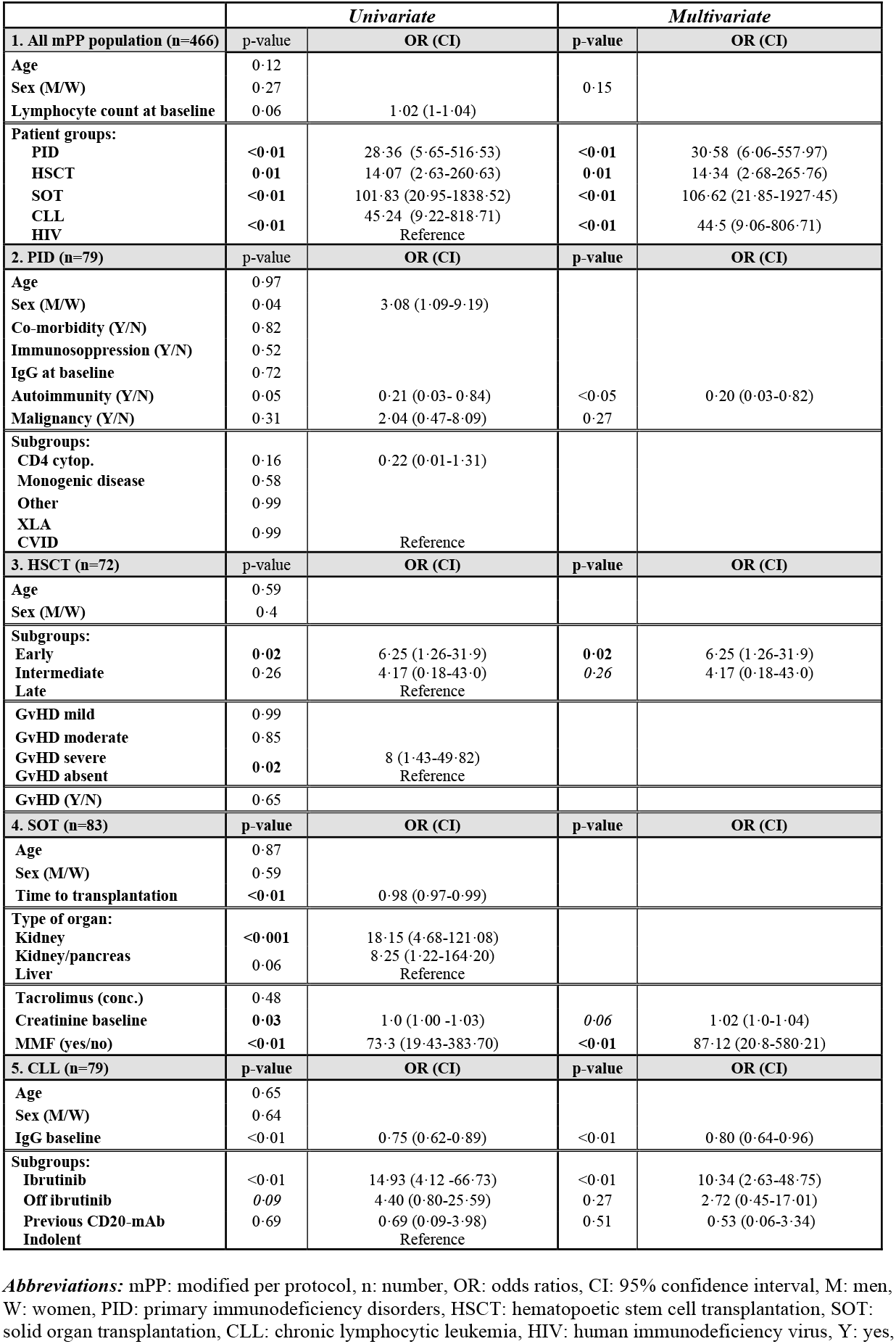
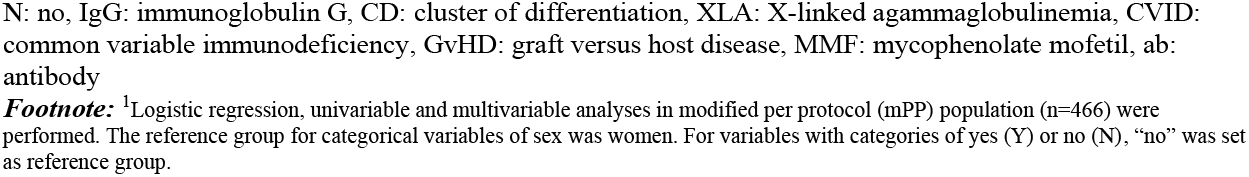
Analysis of factors related to seroconversion failure in the different patient-groups.^1^.

The overall seroconversion rate in the CLL group was 63·3% (p<0.01 compared to controls). Analyzing the subgroups, patients with the lowest seroconversion rate were found in the ongoing ibrutinib (a BTK inhibitor) treatment group (26·9%). The rate doubled in those who had previously been treated with ibrutinib (55·6%). Indolent and patients off long-term chemoimmunotherapy had seroconversion rates >80% (Table 4 and Figure 3A). Treatment with ibrutinib had a negative impact on the likelihood for seroconversion in multivariate analysis. 16/18 patients (88·9%), who had previously (median 13 months; range 7 – 29 months) been treated with anti-CD20 responded. Normal levels of IgG at baseline were positively correlated with seroconversion (Table 5).

The overall seroconversion rate in the PID group was 73·3% (p<0.01 compared to controls). Analyzing the subgroups, patients with common variable immunodeficiency (CVID) had the lowest seroconversion rate (68·3%), followed by patients with monogenic PIDs (77·8%). Patients with low CD4-counts and other PIDs had almost normal seroconversion rates (90·9% and 100%, respectively). As expected, patients with X-linked agammaglobulinemia (XLA) failed to produce any spike specific IgG after vaccination (Table 4 and Figure 3A). The overall seroconversion rate in the HSCT group was 84·7% (p=0.02 compared to controls). Analyzing the subgroups, time after HSCT (<6 months and 6-12 months) significantly influenced the seroconversion compared to healthy controls (Table 4 and Figure 3B). Univariate, but not multivariate analysis, identified severe chronic GvHD as a risk factor for failure to seroconvert (Table 5). Two patients with CD19 CAR T cell treatment failed, as expected, to produce any spike-protein specific IgG after vaccination (Table 4 and Figure 3A).

Finally, the overall seroconversion rate in the HIV group was 98·7% (p =NS compared to controls), with no significant differences in the CD4 cell count subgroups (>300 CD4 cells/µl and <300 CD4 cells/µl, respectively) (Table 4 and Figure 3A).

Additional results on SARS-CoV-2 antibody titres (U/ml) are depicted on a study group level (healthy controls, PID, HIV, HSCT/CAR T, SOT, and CLL) in Figure 2B-D. Generally, significant lower SARS-CoV-2 specific antibody titres were observed in the CLL, SOT and PID group in line with the seroconversion rates (Figure 2B-D). Furthermore, SARS-CoV-2 specific antibody titres varied significantly within different subgroups of the specific patient groups (Figure 3B).

### COVID-19 infections during the study

Twenty-five study subjects (25/539, 4·6%) were found to be seropositive at baseline, among whom two (0·4%) were also RT-PCR positive for SARS-CoV-2. Further description of these patients is provided in the Supplementary Text. The study subjects’ antibody titres are shown in Supplementary Figure 1. Eleven study subjects (2·0%; 5 PID, 3 HSCT, 1 SOT, 2 controls) were diagnosed with COVID-19 between the first and second dose of vaccine. Among the eleven patients, the severity was ≥grade 3 in three patients and severity grade 7 in one patient (scale, see Supplementary Text). Additionally, one patient from the SOT-group, with seroconversion failure at day 35, developed severity grade 2 COVID-19 at 19 days after the second dose.

## Discussion

This study reports the results of a prospective clinical trial evaluating the safety and humoral immune responses following two doses of COVID-19 mRNA BNT162b2 vaccination in five selected groups of immunocompromised patients and healthy controls. The patient groups included were selected to represent different types of primary immunosuppression conditions as well as different secondary immunosuppression states. This readily allows comparisons between specific patient groups and healthy controls. Administration of two consecutive doses, 3 weeks apart, of BNT162b2 was overall safe. The rate of seroconversion was generally lower in immunocompromised patients compared to healthy controls with the lowest responses in the SOT and CLL patient groups. The prospective design of the study furthermore allowed analyses of risk factors for seroconversion failure, in addition to prospective analysis of safety.

SOT patients showed the lowest overall seroconversion with only 43·4% responding. Receiving MMF as a part of the immunosuppressive treatment was strongly associated with low seroconversion, which is in line with previous studies ^10,12,13^. A recently published report found that a third vaccine dose increased the seroconversion rate in SOT patients from 40% to 68% ^14^. This, however, still leaves almost one third of SOT patients without a serological response. As the present results indicate, a possible strategy might be to temporarily discontinue MMF to increase the chance of a vaccine response. This intervention must be weighed against the risk of development of donor specific antibodies or even T-cell mediated rejection of the graft.

The first reports on COVID-19 vaccination in CLL patients found that only 39·5% of included patients seroconverted ^15^. The corresponding rate in our clinical trial was 63·3%. Seroconversion was generally low (26·9%) in patients with ongoing ibrutinib therapy, but nearly doubled in those who had stopped/paused this therapy, in line with previous reports ^15,16^. In contrast, >80% of the patients who had indolent CLL or were long-term off anti-CD20 based chemoimmunotherapy responded to the vaccine. Previous anti-CD20 therapy has been associated with poor responses to vaccines. In the present study, however, most patients responded after a median time of 13 months between anti-CD20 therapy and vaccination. Hence, actions may be required, particularly in those who are on treatment with ibrutinib where temporary cessation of ibrutinib-treatment before vaccination could be warranted. With respect to patients with PID, a low seroconversion rate was found in patients with CVID. Interestingly, all but one of the patients with idiopathic CD4 cytopenia seroconverted. In addition, a patient with hypomorphic SCID due to a mutation affecting the *Artemis* gene and a patient with a *CARD11*-mutation did not respond to vaccination, supporting the importance of these genes for antibody responses ^17,18^. The results are in line with a previous study in which seroconversion was observed in 18/26 (69·2%) PID patients after vaccination with BNT162b2 ^8^. Overall, we observed that most PID-patients responded to vaccination and the number of AEs was low.

In HSCT patients, the results are concordant with studies of other vaccines. Some of the present findings are also similar to other reports of COVID-19 vaccines in this patient group. Time after HSCT had a significant impact on the likelihood of seroconversion similar to findings in other studies ^19-21^. However, it was observed that severity of chronic GvHD impacted negatively on seroconversion in univariate analysis. Seroconversion failure was furthermore found to be associated with ongoing second line treatments for chronic GvHD, such as ruxolitinib and photophoresis, and administration of anti-CD20 therapy given several months prior to vaccination. An effect of the severity of chronic GvHD has not been reported previously but is not unexpected considering what has been observed for other vaccines. None of the two assessable patients receiving CD19 CAR T cell therapy seroconverted, likely due to the persistent depletion of B cells after successful therapy.

People living with HIV responded well to the vaccine, with high seroconversion rates and antibody titres regardless of low (<300 cells/µl) or high (>300 cells/µl) CD4 counts. These results are in line with recent reports that demonstrated robust humoral BNT162b2 vaccination response in this group ^11,22,23^. However, the durability of the antibody response in PLWH will be important to follow since, despite effective antiviral therapy, full immune reconstitution is not achieved in many PLWH. These individuals can have diminished or less durable response to vaccination, which is particularly relevant to monitor in those with low CD4 cell-counts ^24-26^.

This is to our knowledge the first prospective, clinical trial performed in several immunocompromised patient groups allowing careful assessment of safety. Reactogenicity was comparable to previous reports ^5^, and other AE were also generally mild. However, a few immune activation phenomena were observed, such as four cases of GvHD among the HSCT patients. Similarly, Ali et al. reported recently in a retrospective study that 9·7% of HSCT patients developed new chronic GvHD and 3·5% experienced worsened chronic GvHD after vaccination with mRNA vaccines ^27^. Moreover, Ram et al. reported in a prospective cohort study three cases of worsened GvHD (5%) after each dose of BNT162b2 vaccine among 66 allogenic HSCT recipients ^20^. Of note, the traditional adjuvanted pandemic H1N1 influenza vaccine has also been reported to aggravate chronic GvHD ^28^. Taken together, these observations indicate the necessity for careful monitoring and evaluation in future prospective studies and clinical routine. One case of SUSAR with progressive respiratory failure and fatal outcome occurred. This case will need further evaluation.

It is possible that mRNA-vaccines, by virtue of their potent immunogenicity, may precipitate dysfunctional immune-responses in particularly vulnerable patients and/or patient groups. As would be expected in a large clinical trial comprising of more than 500 individuals during the third wave of COVID-19 infection in Sweden, a few COVID-19 cases were documented during the study. In this respect, the present study was not powered to evaluate a potentially protective effect on the number and severity of COVID-19 cases.

A particular strength of the present study is the clinical trial setting with careful prospective safety evaluation. In addition, the study comprises a relatively large participant number, with *a priori* defined monitoring and analyses of the data. The study clearly shows that not all patient groups have the same risk for poor response to COVID-19 vaccination. For example, HSCT patients at a late stage after transplantation and without chronic GvHD responded well to two doses of vaccine. It is unknown, however, whether the duration of immunity will be similar to healthy controls, which requires further studies with a longer follow-up time. In contrast, we also identified subgroups of patients responding poorly, or very poorly, to vaccination. Some of these risk factors have been previously identified, such as ibrutinib in CLL patients and the use of MMF in SOT patients and such patients might benefit from a 3^rd^ dose of vaccine.

There are several limitations of this study. The trial had an open-label and non-randomized design. However, since the vaccine is approved and recommended by the Public Health Agency of Sweden, it was considered unethical to allocate patients to a non-treatment group. Furthermore, we did not pre-screen for SARS-CoV-2 antibodies. The 4·6% rate of seropositive cases at baseline was somewhat high, given the general recommendation of self-isolation for these patients. However, due to high prevalence of SARS-CoV-2 infection in the Stockholm region at the time of the study, the result should reflect the real-life situation. Finally, we did not include other immunological responses, such as T cell responses, in the predefined primary and secondary endpoints. There is a wide spectrum of immunosuppressive disorders and we studied only some of these. This study may, however, serve as a proof-of-concept study to analyze the impact of specific immunosuppression on the seroconversion rate in some patient groups.

The results presented here show that many immunocompromised patients can respond to two doses of BNT162b2a vaccine against COVID-19. However, substantial proportions of these patients respond poorly and may therefore be in need of additional doses to boost the humoral immune response. Indeed, recent reports have shown that immunocompromised SOT-patients with negative antibodies after two doses of mRNA vaccine can respond to a third dose with production of specific antibodies ^14,29^. A third dose of COVID-19 vaccine has just (August 26, 2021) been recommended to immunosuppressed patients by the Swedish Public Health Authority. Similar recommendations have recently also been introduced by other national authorities such as the US CDC and the corresponding French Authority.

In conclusion, this prospective clinical trial showed that the mRNA BNT162b2 vaccine is safe to administer to immunocompromised patients. However, the rate of seroconversion is substantially lower compared to healthy controls, with a wide range of seroconversion rates and titres within the patient groups and subgroups at risk. This knowledge can form the basis for individually adapted vaccination schedules. This might require specific vaccination strategies in different groups of immunosuppressed patients such as subsequent vaccinations for boost, pausing of concomitant immunosuppression, and/or in some cases pre-interventional vaccination.

## Supporting information

Suppl files (tables + COVAXID collaborator group) + suppl figure 1

## Data Availability

Data sharing
Data will be submitted to European Union Drug Regulating Authorities Clinical Trials Database (EudraCT). The full clinical study protocol is available via the SciLifeLab Data Repository (English version: doi:10.17044/scilifelab.15059364; Swedish version doi: 10.17044/scilifelab.15059355). Anonymous data displayed in the manuscript will be made available upon request to the corresponding author following publication of the present article. Data will be made available in a form not deviating from what is accepted by local regulatory authorities with respect to handling of patient data, and in adherence of the policies of the Karolinska University Hospital and Karolinska Institutet.

## Contributors

PB, LH, SM, PN, PC, GS, AÖ, CIES, PN, SH, KL, GL, MSC, MB, HGL, PL and SA contributed to conceptualization, funding acquisition and discussion of data. PL and SA wrote the clinical trial protocol. PB, LH, SM, PN, GS and SA conducted investigation with role of primary investigator for each study participant’s group, recruited study participants and conducted management of participants during the trial. OB, LH, AÖ, LB, JV, EWB, ACN, AT, and AN conducted investigation through recruitment of the study participants and conducted management of participants during the trial. PB, PN, OB, LH, SM, PL, and SA conducted project administration, had access to data, and wrote the original draft. PC contributed to project administration through planning and coordinating the samples, investigation of data collection, and visualization. DW, ACG, and MA contributed to investigation through sample processing. PC, PL, and SA verified the underlying data and contributed to data curation. GB and SMu contributed to investigation through sample analyses. DV contributed to statistical part of planning the study, writing the study protocol, performing formal analysis, and writing original draft. HGL contributed to project administration, resources, and supervision. SA contributed with overall supervision of the trial. All authors reviewed and edited revisions of the manuscript and had final responsibility for the decision to submit for publication.

## Data sharing

Data will be submitted to European Union Drug Regulating Authorities Clinical Trials Database (EudraCT). The full clinical study protocol is available via the SciLifeLab Data Repository (English version: doi:10.17044/scilifelab.15059364; Swedish version doi: 10.17044/scilifelab.15059355). Anonymous data displayed in the manuscript will be made available upon request to the corresponding author following publication of the present article. Data will be made available in a form not deviating from what is accepted by local regulatory authorities with respect to handling of patient data, and in adherence of the policies of the Karolinska University Hospital and Karolinska Institutet.

## Acknowledgements

This study has been funded by the Knut and Alice Wallenberg Foundation, Nordstjernan AB, Region Stockholm (Clinical research position-, ALF-, and CIMED-grants), the Swedish Research Council, a graduate student fellowship from Karolinska Institutet. In addition, the Swedish patient organizations of Primary Immunodeficiencies (PIO) and Hematology (Swedish Blood Cancer Foundation) provided grants. Neither of funders had any role in study design, collection, analysis, interpretation of data or writing of the report.

We thank all the participants enrolled in this study, and Karolinska University Hospital, the sponsor of this clinical trial for making this study feasible. We would like to thank all the research and clinical staff at Karolinska University Hospital, especially research/clinical nurses Linn Wursé, Cecilia Lång, Anna Löwhagen Welander, Susanne Hansen, Douglas Carrick, Katarina Stigsäter, Susanne Cederberg, Annika Olsson, Ingrid Andrén, Margareta Gustafsson, Karin Fransson, Karin Linderståhl, Sonja Sönnert Husa, Kirsti Niemalä, Begüm Eker, Eva Martell, Helena Pettersson, Charlotta Hausmann, Maria Abramsson and Ruza Milosavljevic for their hard work in contributing to the study and data collection. We thank dr. Lena Dillner for support and Sara Roth de Albuquerque for administrative help. We thank dr. Jan Albert from Clinical Microbiology, Karolinska University Hospital and Elisa Pin from SciLifeLab, KTH Royal Institutet of Technology for fruitful discussions, dr. Felicia Hagström for contributing to inclusion of patients, Mats Hellström for creating eCRF database and data withdrawal. We would also like to thank Karolinska Trial Alliance, especially Maria Persson, Ingalill Reinholdsson and Maria Fernström for contributing to work for application to authorities and for monitoring of the study.

## Figure legends

**Figure 2.** Seroconversion and antibody titres per patient group and in healthy controls. A) Seroconversion in the five immunocompromised groups and control group defined as ≥ 0·8 U/ml assessed in the modified per protocol (mPP) population. B) Median SARS-CoV-2 specific antibody titres in the five immunocompromised groups and control group. C) Median (CI 95%) SARS-CoV-2 specific antibody titres at day 35 in individuals who seroconverted. D) Individual antibody dynamics (black thin lines) with median interquartile range (IQR) (coloured thick lines) for each respective group. X-axis: days after first vaccination if not else noted.

**Figure 3.** Seroconversion and antibody titres in subgroups of the specific patient groups. A) Seroconversion in the specific subgroups defined as ≥ 0·8 U/ml in the modified per protocol (mPP) population (see right column for subgroup classification). B) Individual SARS-CoV-2 specific antibody titres for each timepoint in the respective subgroups. Dotted lines represent upper (25,000 U/ml) and lower (0·4 U/ml) limits of detection. Dashed line represents seroconversion threshold of 0·8 U/ml.

**Supplementary Figure 1**. Seroconversion and antibody titres in all patients according to intention to treat (ITT). A) SARS-CoV-2 specific antibody titres ≥ 0·8 U/ml for each time point in the five immunocompromised groups and control group. B) Dynamics of SARS-CoV-2 specific antibody titres for all patients who received at least dose 1. C) Dynamics of SARS-CoV-2 specific antibody titres for healthy controls and all patients in each respective group who received at least dose 1. Seroconversion in patients before receiving dose 1 (red), in patients who had not received dose 2 (blue), or where baseline samples or day 35 samples were missing.

