## Supplementary material for "Safety and efficacy of the mRNA BNT162b2 vaccine against SARS-CoV-2 in five groups of immunocompromised patients and healthy controls in a prospective open-label clinical trial": Suppl files (tables + COVAXID collaborator group) + suppl figure 1

1. Full detailed description of statistical analyses.
2. Grades of COVID-19 severity.
3. Antibody titres after vaccination with BNT162b2 in individuals positive for SARS-CoV-2 antibodies at baseline.
4. Antibody test
5. Supplementary Table 1: Reactogenicity after vaccine doses 1 and 2.
6. Supplementary Table 2: Adverse events after vaccine doses 1 and 2.
7. Supplementary Table 3: Proportion of patients with seroconversion after two doses of BNT162b2 vaccination in per protocol population.
8. Supplementary figure 1. Seroconversion and antibody titers in all patients according to intention to treat (ITT).
9. COVAXID Collaborator group
10. Supplementary References.

### 1. Full detailed description of statistical analyses

Safety analyses included all the participants who received at least one dose of BNT162b2, in intention-to-treat (ITT) analyses. Reactogenicity, adverse events (AE), severe adverse events (SAE) and suspected unexpected serious adverse reaction (SUSAR) after first and second vaccine dose, respectively, were assessed.
Analysis of primary efficacy endpoint with seroconversion included per protocol (PP) analysis, with participants who received two doses of BNT162b2 with estimation of the proportion of participants (95% CI) with positive SARS-CoV-2 antibody tests at day 35. Those with no available sample at day 0 and 35, or positive SARS-CoV-2 PCR/antibody tests at baseline, were excluded in the PP analysis. Analysis of primary efficacy endpoint with seroconversion was also performed on modified PP (mPP) population, with estimation of the proportion of participants with seroconversion (95% CI) at day 35 among participants who received two doses of BNT162b2, who were not seropositive at baseline, and who did not develop COVID-19 during the study (Figure 1). Proportions of seroconversion were compared in patient groups, or prespecified subgroups vs. controls, with estimation of CIs and p-values (Fisher's exact test). Differences in mean antibody titer values at day 35 between groups and subgroups were tested through pairwise comparisons using Wilcoxon rank sum test. Bonferroni correction was applied.

Logistic regression, univariable and multivariable, was used to investigate possible predictive factors for the outcome of negative seroconversion at day 35. Variables with p-values < 0·35 from the univariable analysis were conservatively inserted in the multivariable analysis. The best fitting model was here obtained through stepwise selection. The statistical analyses were performed using R base (R Core Team, 2021).

### 2. Grades of COVID-19 severity

The severity of COVID-19 was assessed with ordinal scale, with scores of 1-8 according to Beigel et al [1].
1, not hospitalized, no limitations of activities.
2, not hospitalized, limitation of activities, home oxygen requirement, or both.
3, hospitalized, not requiring supplemental oxygen and no longer requiring ongoing medical care (used if hospitalization was extended for infection-control reasons).
4, hospitalized, not requiring supplemental oxygen but requiring ongoing medical care (Covid-19–related or other medical conditions).
5, hospitalized, requiring any supplemental oxygen.
6, hospitalized, requiring noninvasive ventilation or use of high-flow oxygen devices.
7, hospitalized, receiving invasive mechanical ventilation or extracorporeal membrane oxygenation (ECMO); and 8, death.

### 3. Antibody titres after vaccination with BNT162b2 in individuals positive for SARS-CoV-2 antibodies at baseline

All 25 individuals with positive SARS-CoV-2 antibodies at baseline received two doses of BNT162b2 (the antibody tests were analyzed after day 35, hence no information of seropositivity was available at the start of the study). Severe adverse events (SAE) were noted in one of these individuals, due to underlying liver failure and primary immune deficiency diagnosis. This event was judged to not be related to the vaccination, according to the primary investigator. Generally, individuals with positive SARS-CoV-2 antibodies reached very high antibody titers (Supplementary Figure 1), with 15 having a titer of > 25,000 U/mL.

### 4. Antibody test

Serum samples were tested using the commercial, quantitative Roche Elecsys anti-SARS-CoV-2 S enzyme immunoassay, which measures total antibodies to the SARS-CoV-2 S receptor-binding domain (RBD) protein, the target of the mRNA vaccines. Results range from <0.4 to >250 U/mL with the positive cut-off defined as >0.79 U/mL. According to the manufacturer, the overall clinical specificity was 99.98% (n=5991) and sensitivity was 98.8 % for samples taken ≥14 days after positive PCR result (n=1423). In an independent assessment, the highest sensitivity (84.0%, n=50) was observed at 15 to 30 days post-PCR positivity and an assay specificity of 100% (n=32) was reported. The assay has also been validated against the first WHO-standard for Anti-SARS-CoV-2 immunoglobulin (NIBSC 20/136) [2]. Our individual assessment of the assay resulted in a specificity of 100% (n=80, collected from patients in 2019) and a sensitivity of 100% at ≥18 – 40 days post-PCR positivity (n=37). During validation an intra-assay CV (coefficient of variability) of 0.8% and an inter-assay of CV 0,9% was observed across 3 days and using one reagent lot. Since introduction of the assay in our laboratory in February 2021 inter-assay variation has been continuously monitored and showed satisfactory values (≤ 8.7%).

### 5. **Supplementary Table 1:** Reactogenicity after vaccine doses 1 and 2.

**After Dose 1**

| Any reaction (grade 1 – 4) | Patients no. | % | Controls no. | % |
|---|---|---|---|---|
| **Yes** | 368 | 81·9 | 71 | 78·9 |
| **No** | 67 | | 26 | |
| **Missing** | 14 | | 3 | |
| **Total patients vaccinated** | 449 | | 90 | |
| | N=435 | | N = 87 | |
| **Local (pain, redness, swollen) grade 2 – 4** | 85 | 20.0 | 15 | 16.7 |
| **Systemic (fever, tiredness, chills, headache) grade 2-4** | 93 | 18·9 | 21 | 23·3 |
| **Gastrointestinal grade 2-4** | | | | |
| **Diarrhea** | 13 | 2.9 | 0 | 0 |
| **Vomiting** | 2 | 0·4 | 1 | 1·1 |
| **Joint or muscle pain grade 2 – 4** | 29 | 6·4 | 2 | 2·2 |

**After Dose 2**

| Any reaction (grade 1 – 4) | Patients no. | % | Controls no. | % |
|---|---|---|---|---|
| **Yes** | 333 | 76·3 | 74 | 84.1 |
| **No** | 91 | | 13 | |
| **Missing** | 14 | | 1 | |
| **Did not receive dose 2** | 13 | | 2 | |
| **Total patients vaccinated** | 436 | | 88 | |
| | N=422 | | N=87 | |
| **Local (pain, redness, swollen) grade 2 – 4** | 98 | 22·4 | 18 | 20·.4 |
| **Systemic (fever, tiredness, chills, headache) grade 2-4** | 86 | 19·7 | 29 | 32·9 |
| **Gastrointestinal grade 2-4** | | | | |
| **Diarrhea** | 13 | 3·0 | 2 | 2·2 |
| **Vomiting** | 2 | 0·4 | 1 | 1·1 |
| **Joint or muscle pain grade 2 – 4** | 48 | 11·0 | 8 | 9·1 |

**6. Supplementary Table 2:** Adverse events (non-severe adverse events, SAEs) after vaccine doses 1 and 2.

**After Dose 1**

| | Patients | | Controls |
|---|---|---|---|
| **No. of Adverse Events (AE)** | 74 | | 6 |
| **No. of individuals experiencing AE** | 57 | | 5 |
| **Grades 2-4 CTCAE** | 21 | | 0 |
| **Probably or possibly related** | 7 | | NA |
| **Unlikely to be related** | 14 | | NA |
| **Classification of possibly/probably related AE grades 2-4** | | | |
| Thrombocytopenia | 1 | | |
| Neutropenia | 1 | | |
| Increase in liver function tests | 2 | | |
| Increase in s-creatinine | 1 | | |
| Musculoskeletal | 1 | | |
| Skin/mucous membranes | 0 | | |
| Cardiovascular | 3 | | |
| Mental/neurological | 1 | | |
| Gastrointestinal | 3 | | |
| Infections | 8 | | |
| Pulmonary | 0 | | |

**After Dose 2**

| | Patients | | Controls |
|---|---|---|---|
| **No. of Adverse Events** | 74 | | 1 |
| **No. of individuals experiencing AE** | 58 | | 1 |
| **Grades 2 – 4 CTCAE** | 15 | | 0 |
| **Probably or possibly related** | 6 | | NA |
| **Unlikely to be related** | 9 | | NA |
| **Classification of possibly/probably related AE grades 2-4** | | | |
| Thrombocytopenia | 0 | | |
| Neutropenia | 2 | | |
| Increase in liver function tests | 1 | | |
| Increase in s-creatinine | 0 | | |
| Musculoskeletal | 2 | | |
| Skin/mucous membranes | 2 | | |
| Cardiovascular | 1 | | |
| Mental/neurological | 1 | | |
| Gastrointestinal | 1 | | |
| Infections | 4 | | |
| Pulmonary | 1 | | |

***Abbreviations:*** AE: adverse events, CTCAE: Common Terminology Criteria for Adverse Events , NA: not applicable.

**7. Supplementary Table 3:** Number and proportion of patients with seroconversion after two doses of BNT162b2 vaccination; i.e., seropositive at day 35, in per protocol (PP) population[1].

| | All patients | PID | HIV | HSCT/ CAR-T | SOT | CLL | CTRL |
|---|---|---|---|---|---|---|---|
| **Seroconverted** | 282 | 55 | 78 | 63 | 36 | 50 | 78 |
| **Seronegative** | 108 | 20 | 1 | 11 | 47 | 29 | 0 |
| **Total** | 390 | 75 | 79 | 74 | 83 | 79 | 78 |
| **Seroconversion (CI) (%), P-value** | 72.3 (67.6 – 76.7) P<0.001 | 73.3 (63.3-83.3) P<0.01 | 98.7 (96.3-100) P=1 | 84.7 (76.4-93) P<0.01 | 43.4 (32.7-54) P<0.01 | 63.3 (52.7-73.9) P<0.01 | 100 |

***Abbreviations:*** PID: primary immunodeficiency disorders, HIV: human immunodeficiency virus, HSCT: hematopoetic stem cell transplantation, SOT: solid organ transplantation, CLL: chronic lymphocytic leukemia, CTRL: healthy controls, CI: 95% confidence interval.
***Footnote****:* [1]P-values of the differences vs. healthy controls were calculated.

### 8. COVAXID Collaborator group (to be indexed in PubMed)

**Xinling Xu**, MD, Department of Infectious Diseases, Karolinska University Hospital, Stockholm, Sweden

**Katarina Westling**, MD, Department of Infectious Diseases, Karolinska University Hospital, Stockholm, Sweden; Department of medicine, Div of Infectious Diseases and Dermatology Karolinska Institutet, Sweden

**Lina Lindström-Älgå**, MD, Department of Transplantation, Karolinska University Hospital, Stockholm, Sweden; Department of Clinical Science, Intervention and Technology, Karolinska Institutet, Sweden

**Greg Nowak**, MD, Department of Transplantation, Karolinska University Hospital, Stockholm, Sweden; Department of Clinical Science, Intervention and Technology, Karolinska Institutet. Sweden

**Christina Villard**, MD, Department of Transplantation, Karolinska University Hospital, Stockholm, Sweden; Department of Clinical Science, Intervention and Technology, Karolinska Institutet, Sweden.

**Andreas Björklund**, MD, Department of Allogeneic Stem Cell Transplantation and Cellular Therapy, Karolinska University Hospital, Sweden

**Marzia Palma**, MD, PhD, Department of Hematology, Karolinska University Hospital/Department of Oncology-Pathology, Karolinska Institutet, Stockholm, Sweden

**Rula Zain**, PhD, Department of Laboratory Medicine, Karolinska Institutet, Sweden.

**Katie Healy**, PhD, Department of Dental Medicine, Karolinska Institutet, Sweden

**Fredrika Hellgren**, MSc, Department of Medicine Solna, Karolinska Institutet, Sweden

#### Contribution of the collaborators

XX, KW, LLÄ, GN, MP and CV have contributed to investigation through recruitment of the study participants and conducted management of participants during the trial. RZL have contributed to discussion regarding conceptualization and funding acquisition. KH and FH have contributed to investigation through sample processing. All authors reviewed and edited revisions of the manuscript and had final responsibility for the decision to submit for publication.

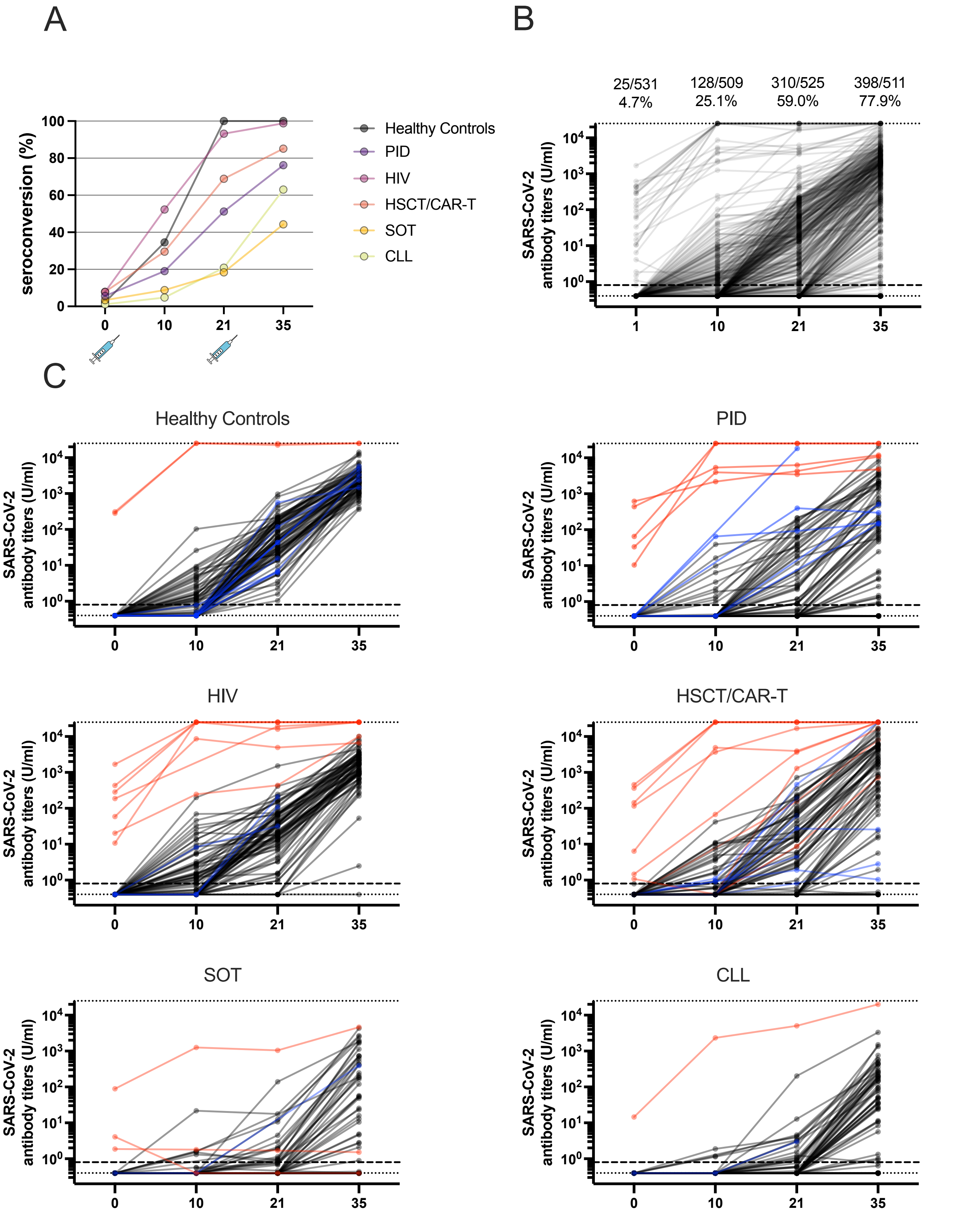

**Supplementary Figure 1.** Seroconversion and antibody titers in all patients accoring to intention to treat (ITT). A) SARS-CoV-2 specific antibody-titers ≥0.8 U/ml for each time point in the 6 cohorts. B) Dynamics of SARS-CoV-2 specific antibody titers for all patients who recieved dose 1. C) Dynamics of SARS-CoV-2 specific antibody titers for all patients in each cohort who recieved dose 1. (red) seroconversion before recieving dose 1, (blue) patients who had not recieve dose 2, no baseline sample or no day 35 sample.
